# Test-adjusted results of mortality for Covid-19 in Germany, USA, UK

**DOI:** 10.1101/2020.11.03.20225268

**Authors:** Jürgen Mimkes, Rainer Janssen

## Abstract

In a disease, where all infected persons show symptoms, it is reasonable to calculate mortality by case to fatality rate CFR. Deaths follow infections by a certain time lag. However, in the Covid-19 pandemic many infectious patients show no or hardly any symptoms. The reported infections and deaths do not run parallel, but diverge with the volume of tests.

Our investigations for Germany, USA and UK indicate that deaths do not follow the number of infections, but the positive rate of tests, multiplied by a constant factor F and shifted by about two weeks. These test adjusted results of mortality allow for the estimation of the number of deaths of Covid-19 about two weeks ahead, even in a sharply rising state of the pandemic. This gives medical authorities two weeks of time to plan for resources.

## Introduction

Mortality is the most important indicator of a pandemic and scientists all over the world are estimating mortality from Covid-19. A good introduction has been given in a scientific brief by the World Health Organization WHO [1]. In a pandemic, two time series are generally reported: the time series of daily infections (I _k_), and the time series for daily deaths (D _k_), as provided by the Johns-Hopkins-University (JHU) in USA and the Robert-Koch-Institute (RKI) in Germany. A wide range of values has been reported in the literature for the CFR, varying with time and location [2–4]. In this paper we concentrate on mortality in Germany, USA, and UK.

### The RKI data for Germany

The series of daily registered new infections (I _k_) and the series of daily deaths (D _k_) are given in figs. 1 and 2.

**Fig. 1.**
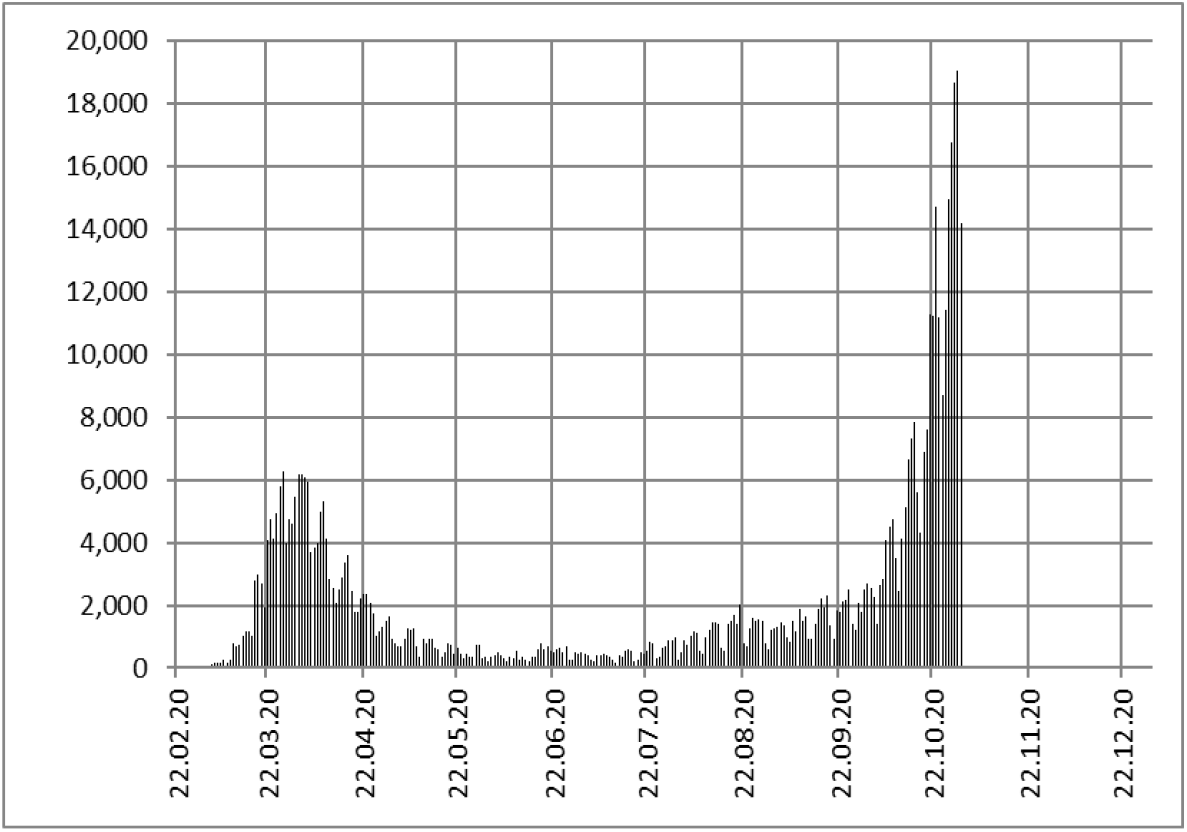
Daily new infections I_k_ in Germany (RKI)

**Fig. 2.**
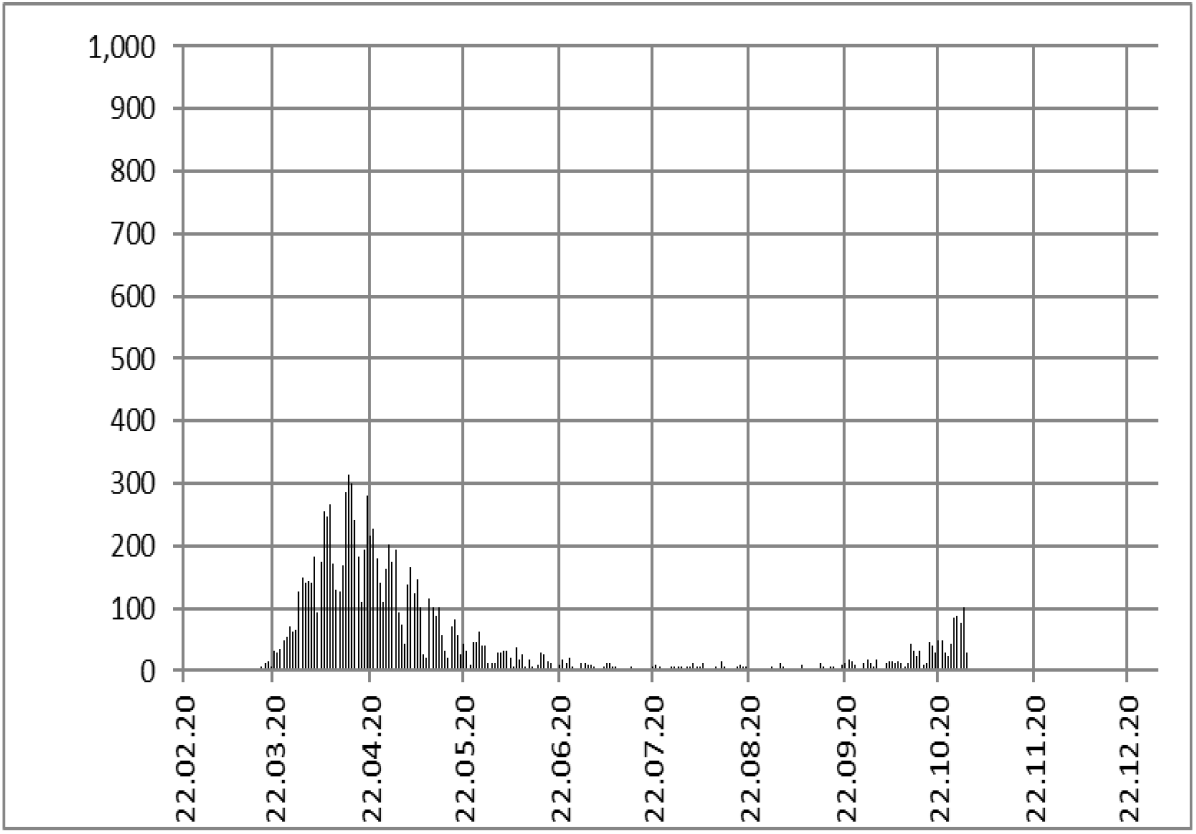
Daily deaths D _k + 13_ in Germany (RKI)

### Calculation of CFR mortality

Normally, in mortality calculations the series of daily deaths (D _k + L_) follows the series of daily infections (I _k_) by a time lag (L) and is smaller by the case fatality ratio CFR,

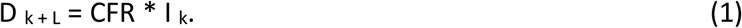

In fig. 2 the maximum of deaths (300) follows the maximum of infected (6000) in fig. 1 about two weeks (L = 13) later, and is smaller by the factor CFR = 300 / 6000 = 0.05. This leads to

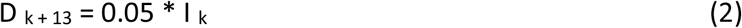

Eq. (2) has been plotted in fig. 3.

**Fig. 3.**
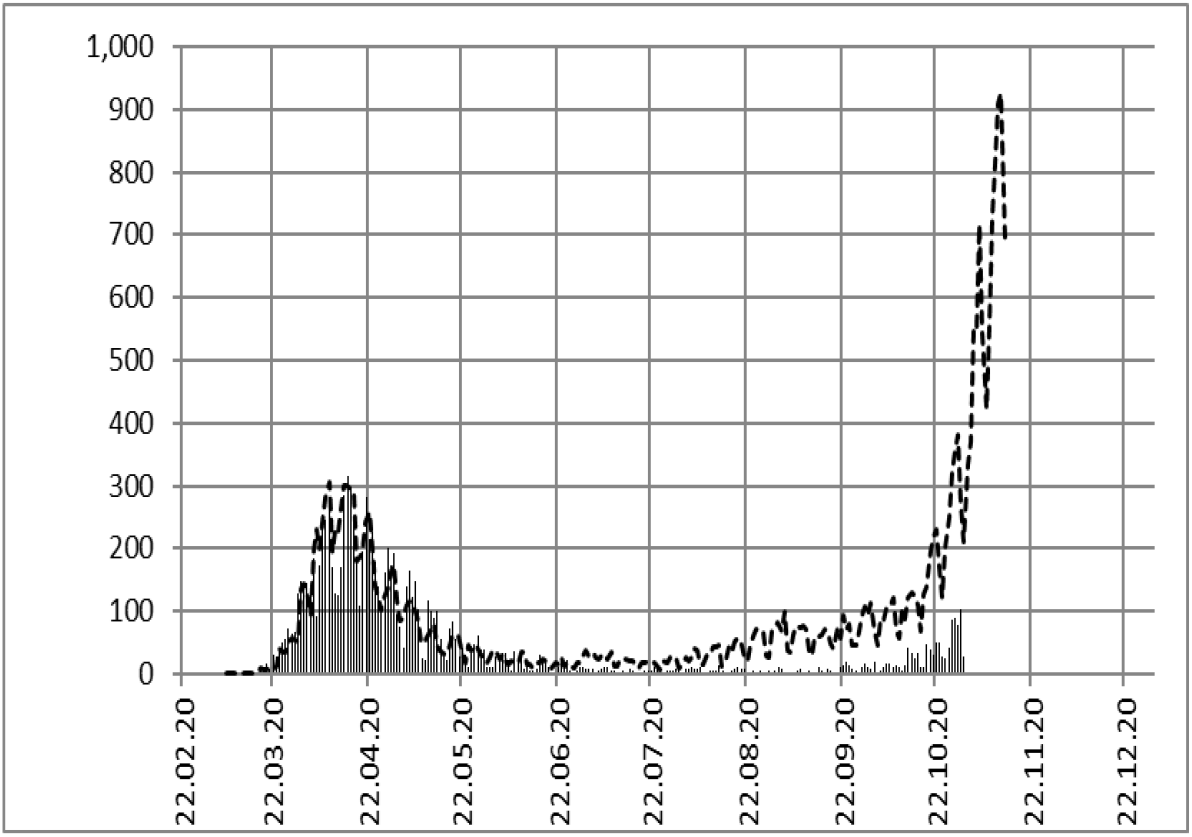
Daily deaths data by RKI (bars) and calculation of D _k + 13_ according to eq. (2) (dotted line).

Data and calculation of CFR mortality in fig. 3 agree for the first Covid-19 wave from March to June, as would be expected. But from July on the calculations of daily deaths drifts away to higher numbers, whereas the actual death data stay close to zero between July and September, indicating only rising numbers in October. As the deaths in fig. 3 do not follow the infections, the CFR is not any more constant, but is time variant, CFR _k_. A similar observation has been made in many countries, as shown for USA and UK, below. In a recent paper the authors [5, 6] proposed to explain the disagreement between calculations (eq. 1) and mortal data in fig. 3 by considering a change in test volume. Indeed, the test volume in fig. 4 in Germany has been increased from July until September by a factor of 3.

**Fig. 4.**
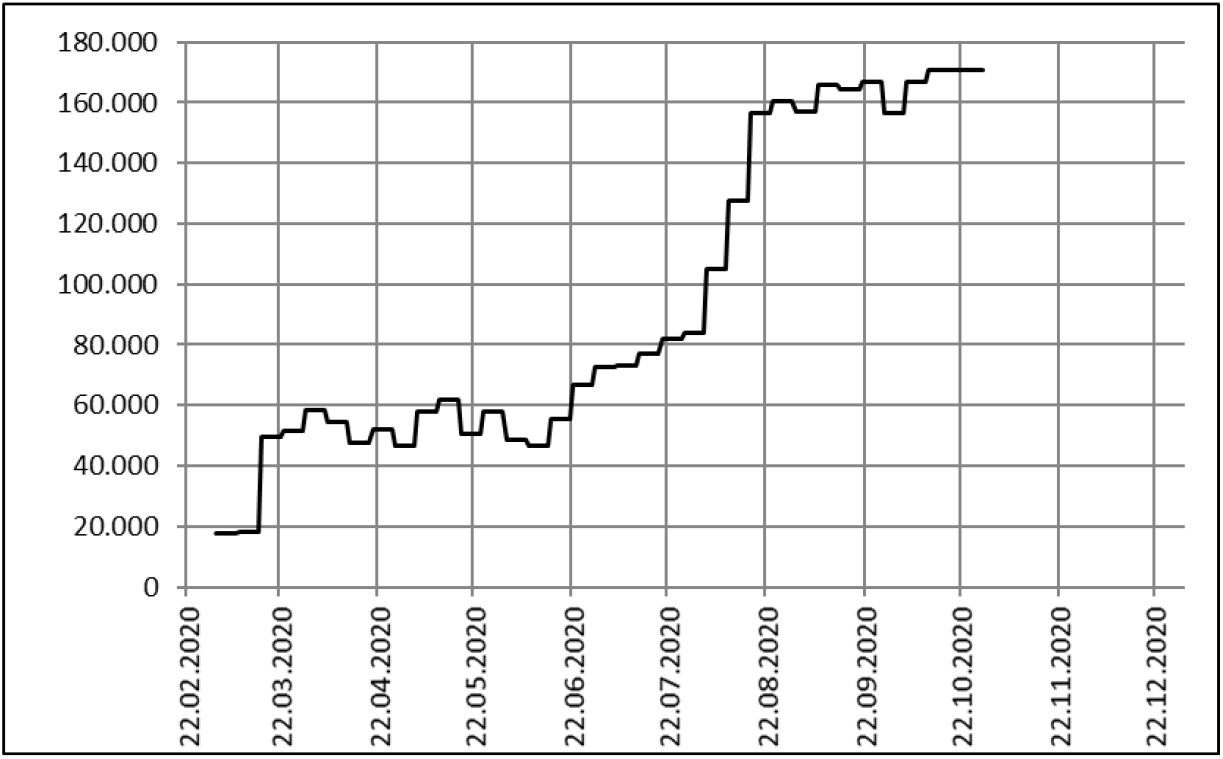
Daily test volume T _k_ (weekly reports by RKI)

For a more detailed analysis, we divide the infection numbers I _k_ by the test volume T _k_. Then we scale the result by a mortality factor F = 2500 to obtain an optimal approximation to the deaths series of the pandemic in fig. 5.

**Fig. 5.**
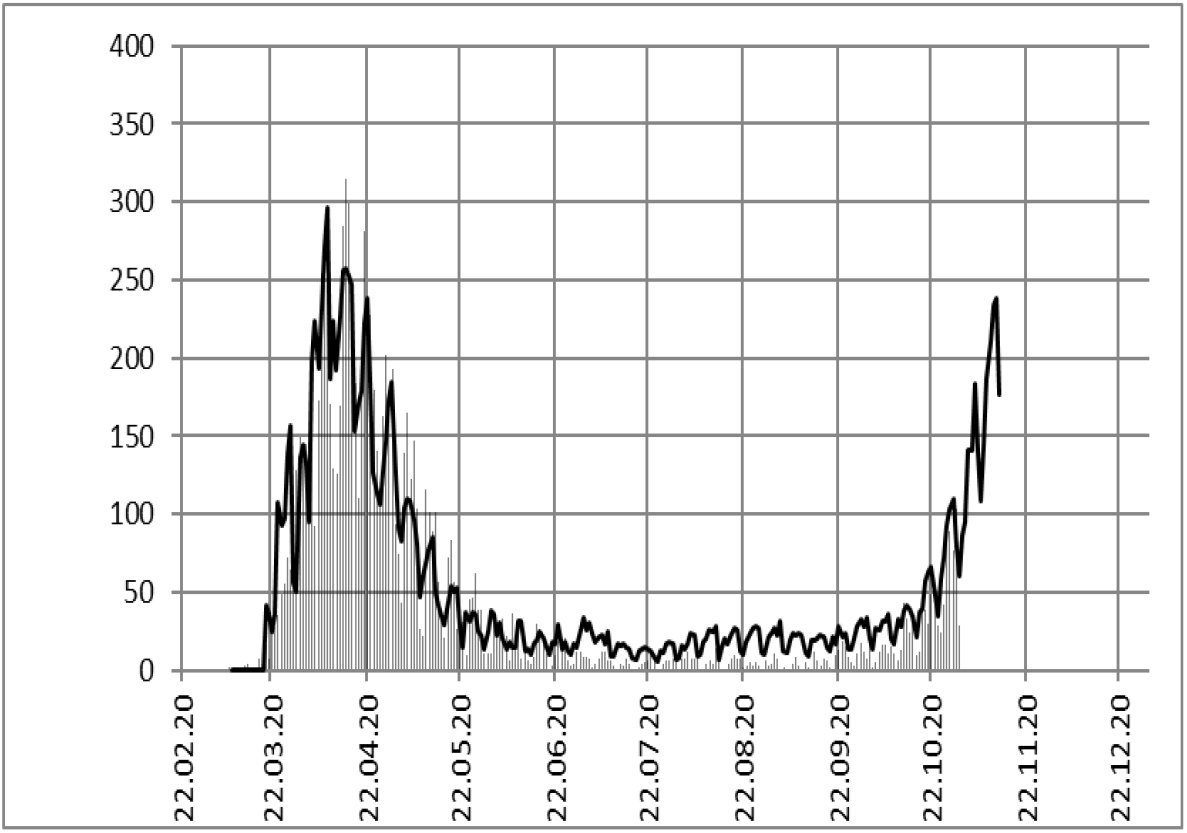
Daily deaths data from RKI (bars) and infection numbers divided by test volume (solid line).

Apparently, in fig. 5 (solid line) the test adjusted infection data (I _k_ / T _k_) are in good agreement with the deaths data (bars) throughout the pandemic,

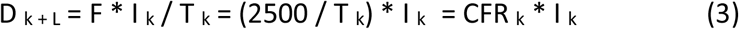

and we obtain a test depending mortality CFR _k_ = 2500 / T _k_.

### A test adjusted approach to mortality

In order to explain the test depending mortality CFR _k_ in eq. 3 and fig. 5, we propose a test adjusted approach to mortality: In diseases where many infectious patients show no or hardly any symptoms, we do not know the total number of infections. But we can determine the daily number of infections I _k, T_ by the positive rate P _k_ of daily infections of a certain daily test volume T _k_,

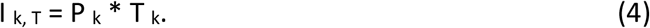

In eq. 3 we may now replace the ratio I _k_ / T _k_ by the positive rate of tests P _k_ of eq. 4,

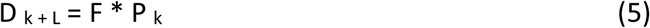

In eq. 5 and fig. 5 the number of daily deaths D _k + L_ follows the daily positive rate P _k_, and not the daily number of infections I _k_ as in eq. 1. The daily numbers of infections and deaths are now represented in eq. 4 and eq. 5 by the daily positive rate P _k_ and the daily test volume T _k_. The daily test volume T _k_ and the daily positive rate of infections P _k_ are given by the Robert-Koch-Institute, in figs. 4 and 6. The time lag L between infection and death is about 13 days in Germany.

**Fig. 6.**
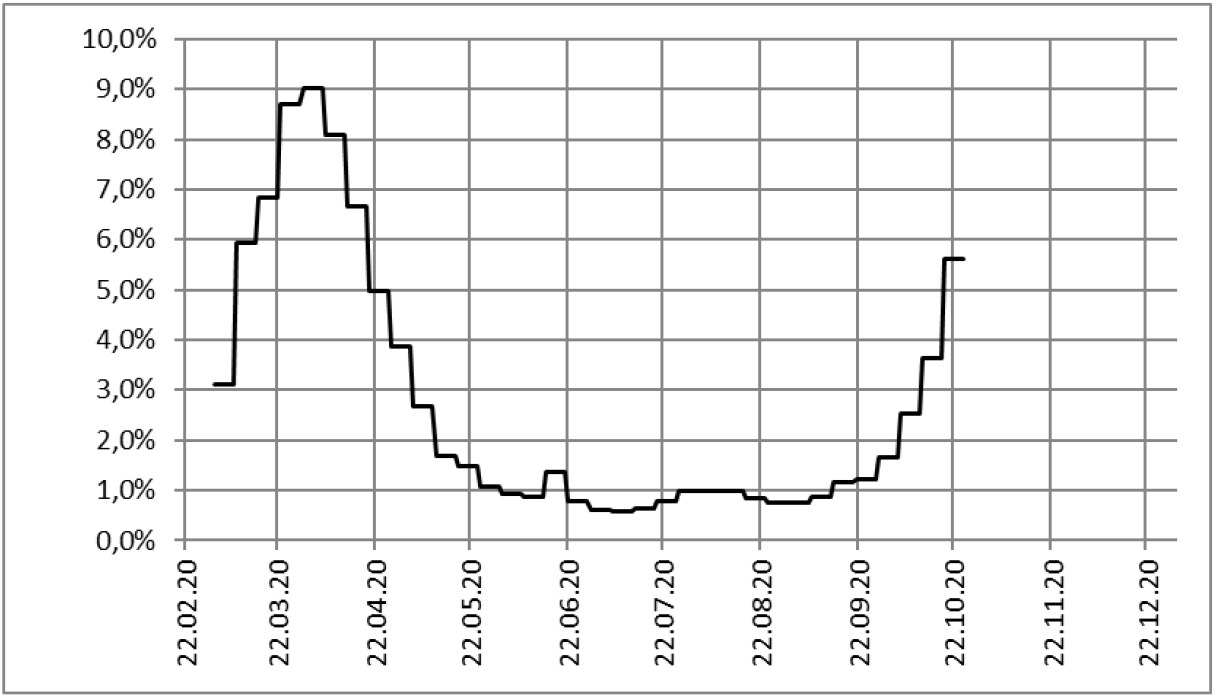
Positive rate P _k_ of daily tests (from weekly reports by RKI)

The relationship between the number of daily deaths and the number of daily infected in eq. (1) is **invalid**, if the daily test volume is not constant. High numbers of infected do not necessarily mean high number of deaths.

*The number of deaths is only determined by the positive rate of daily infections P* _*k*_, *multiplied by a constant mortality factor F*.

This statement does not diminish the danger of Covid-19, but puts it on a scientific basis. Accordingly, the daily positive rate of tests should be published in addition to the presently dominating infection numbers, which are used for other purposes like tracking.

*The daily positive ratio of tests leads to a realistic estimation of the number of deaths two weeks in advance in eqs. 3 or 5 and in fig. 5*.

This gives medical authorities two weeks of time to plan ahead for the numbers of severely ill patients, which are closely related the number of deaths.

### JHU data for USA

For USA, the Johns-Hopkins-University (JHU) provides the series of daily registered new infections (I _k_) and the series of daily deaths (D _k_) in figs. 7 and 8. The daily test series is shown in fig. 9.

**Fig. 7.**
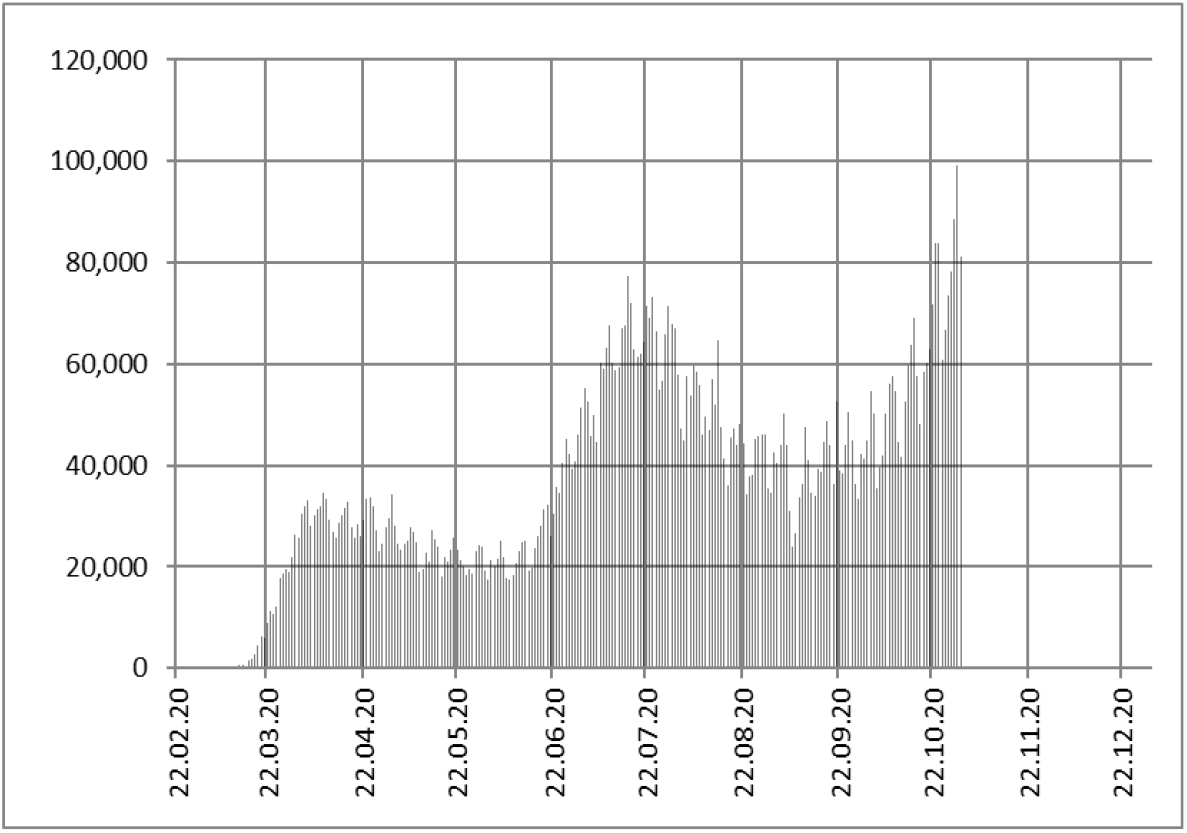
Daily new infections I _k_ in USA (JHU)

**Fig. 8.**
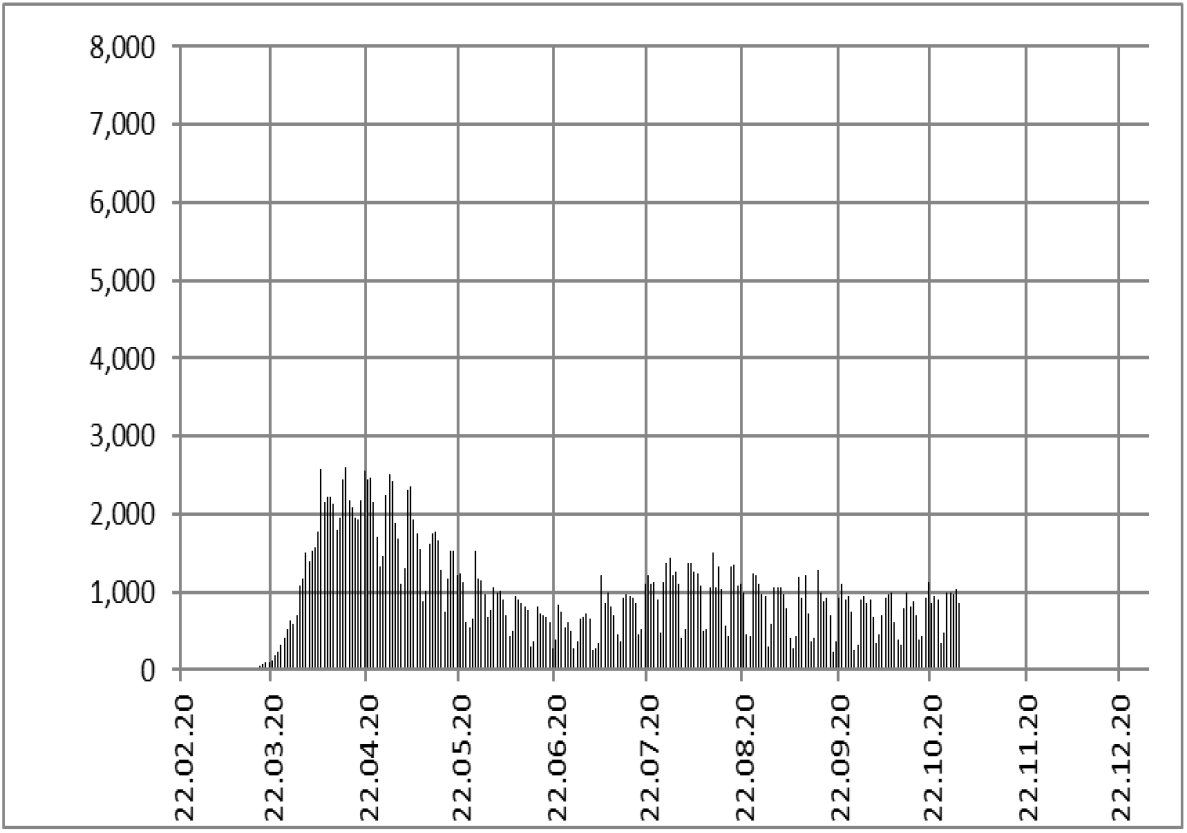
Daily new deaths D _k + 14_ in USA (JHU)

**Fig. 9.**
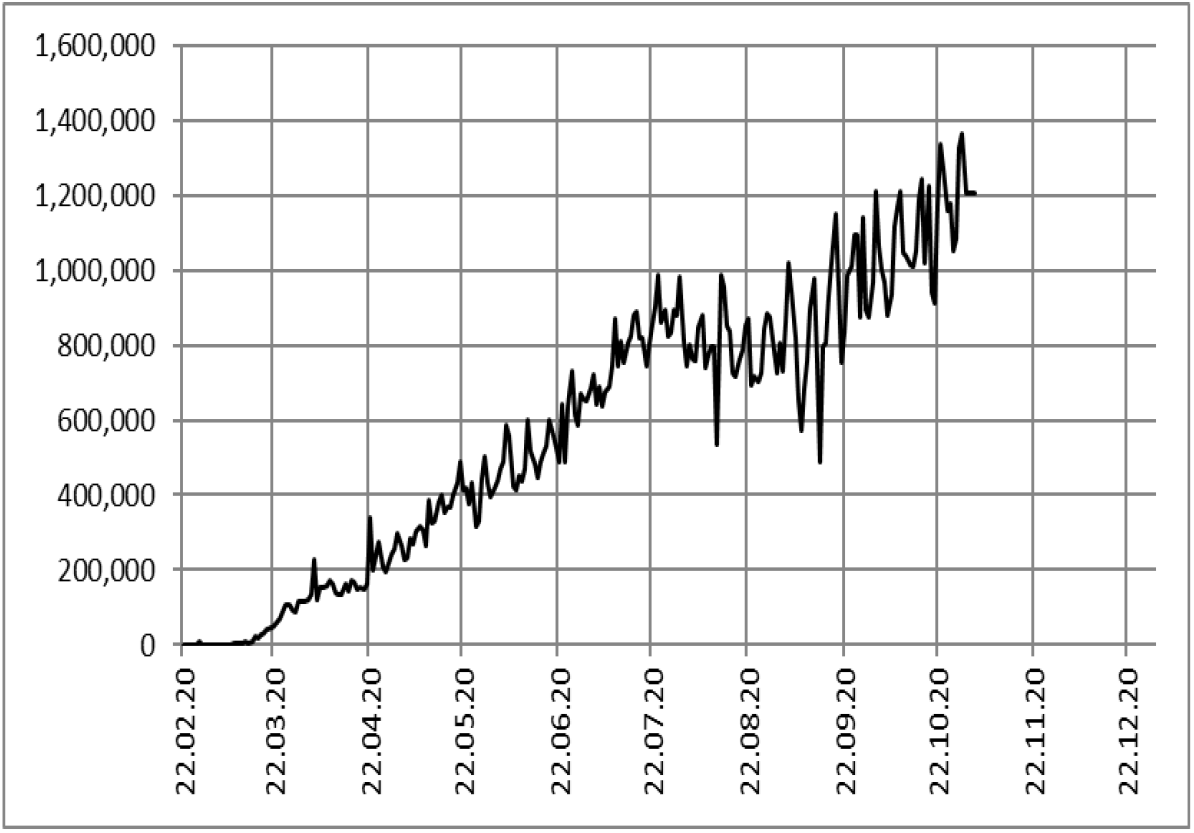
Daily test volume T _k_ in USA (The Covid Tracking Project [7])

Fig. 10 shows the calculation of the series of deaths (D _k + L_) according to eq. 1 by a dashed line and according to fig. 5 by a solid line.

**Fig. 10.**
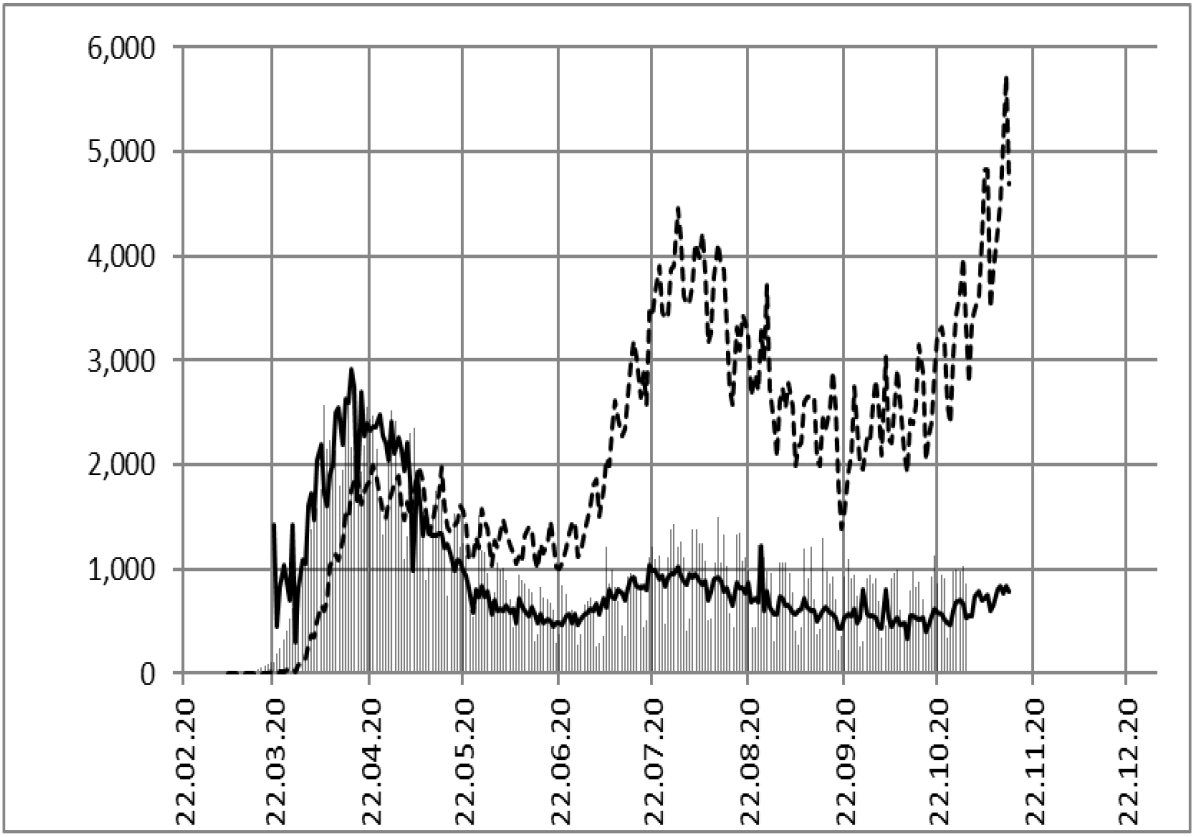
Daily deaths data by JHU (bars) and calculation of D _k + 14_ according to eq. 1 (dashed line) and eq. 5 (solid line).

Fig 10 shows again a good agreement of the test adjusted infection data (I _k_ / T _k_) with the deaths data. The time lag L is about 14 days and the mortality factor about F _USA_ = 11,500. The mortality factor F calculates the number of deaths of a country at a certain positive rate of testing (P _k_), accordingly, the mortality factor will depend on the population of a country if the modalities of testing are comparable.

Since the population in USA (320 million) is larger than the population in Germany (83 million) by a factor of about 4, we would estimate F _USA_ = 4 * F _Ger_ = 10,000. This estimation is of the same order as the result above, and supports the test adjusted approach to mortality of Covid-19 in Germany and other countries.

### JHU data for UK

In UK, the test adjusted infection data (I _k_ / T _k_) also lead to a realistic prediction of the death series of the Covid-19 pandemic (fig. 11). The time lag is about 14 days and the mortality factor F _UK_ about 2,700, deviating slightly from those in USA and Germany, if related to the population.

**Fig. 11.**
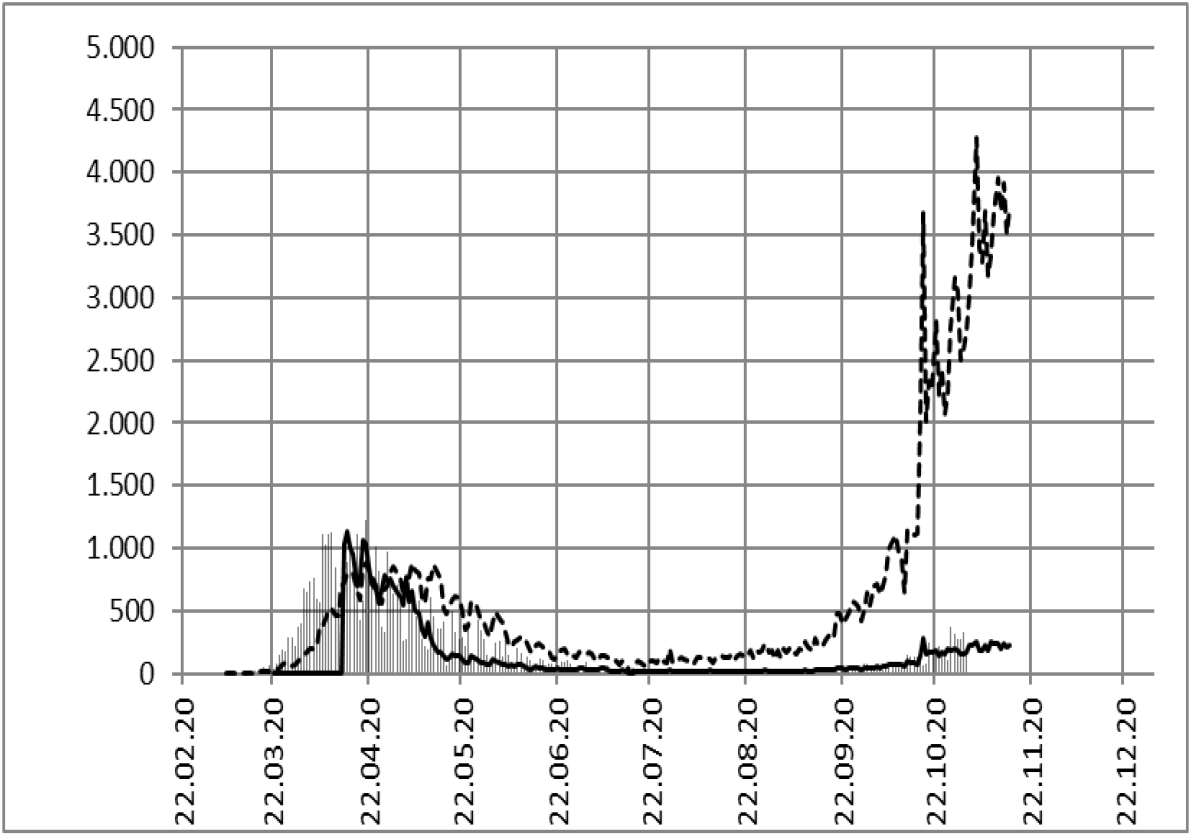
Daily deaths data by JHU (bars) in **UK** and calculation of D _k + 14_ according to eq. 1 (dashed line) and eq. 5 (solid line). Test data since 31.03.20 [8].

## Conclusions

In a pandemic with symptomatic and asymptomatic courses of the disease like in Covid-19, the pure number of infections is not significant for the danger of the pandemic. The indicator for the number of deaths is the positive rate (P _k_) of tests. It is a scale for the number of deaths, and in addition, it is known about two weeks earlier.

## Data Availability

see References

